# A Computational Framework for Intraoperative Pupil Analysis in Cataract Surgery

**DOI:** 10.1101/2024.03.13.24304223

**Authors:** Binh Duong Giap, Karthik Srinivasan, Ossama Mahmoud, Dena Ballouz, Jefferson Lustre, Keely Likosky, Shahzad I. Mian, Bradford L. Tannen, Nambi Nallasamy

**Author notes:** **Corresponding Author:** Nambi Nallasamy, MD Kellogg Eye Center, University of Michigan 1000 Wall St., Ann Arbor, MI 48105, USA. **Disclosures:** The authors declare that they have no known competing financial interests or person relationships that could have appeared to influence the work reported in this paper. **Funding support:** GME Innovations Fund (NN, BT), The Doctors Company Foundation (NN, BT), NIH K12EY022299 (NN), Fogarty/NIH D43TW012027 (NN, KS).

## Abstract

**Purpose:** Pupillary instability is a known risk factor for complications in cataract surgery. This study aims to develop and validate an innovative and reliable computational framework for the automated assessment of pupil morphologic changes during the various phases of cataract surgery.

**Design:** Retrospective surgical video analysis.

**Subjects:** Two hundred forty complete surgical video recordings, among which 190 surgeries were conducted without the use of pupil expansion devices and 50 were performed with the use of a pupil expansion device.

**Methods:** The proposed framework consists of three stages: feature extraction, deep learning (DL)-based anatomy recognition, and obstruction detection/compensation. In the first stage, surgical video frames undergo noise reduction using a tensor-based wavelet feature extraction method. In the second stage, DL-based segmentation models are trained and employed to segment the pupil, limbus, and palpebral fissure. In the third stage, obstructed visualization of the pupil is detected and compensated for using a DL-based algorithm. A dataset of 5,700 intraoperative video frames across 190 cataract surgeries in the BigCat database was collected for validating algorithm performance.

**Main Outcome Measures:** The pupil analysis framework was assessed on the basis of segmentation performance for both obstructed and unobstructed pupils. Classification performance of models utilizing the segmented pupil time series to predict surgeon use of a pupil expansion device was also assessed.

**Results:** An architecture based on the FPN model with VGG16 backbone integrated with the AWTFE feature extraction method demonstrated the highest performance in anatomy segmentation, with Dice coefficient of 96.52%. Incorporation of an obstruction compensation algorithm improved performance further (Dice 96.82%). Downstream analysis of framework output enabled the development of an SVM-based classifier that could predict surgeon usage of a pupil expansion device prior to its placement with 96.67% accuracy and AUC of 99.44%.

**Conclusions:** The experimental results demonstrate that the proposed framework 1) provides high accuracy in pupil analysis compared to human-annotated ground truth, 2) substantially outperforms isolated use of a DL segmentation model, and 3) can enable downstream analytics with clinically valuable predictive capacity.

## INTRODUCTION

Cataract surgery is one of the most commonly performed surgeries worldwide and is essential to addressing preventable blindness. In 2015, there were more than 20 million surgeries performed worldwide, of which 3.6 million cases were in the United States and 4.2 million cases were in the European Union.^1^ Performing cataract surgery requires access to the crystalline lens, which in turn requires adequate dilation and stability of the pupil. The pupil is typically pharmacologically dilated for cataract surgery through the use of medications administered preoperatively and/or intraoperatively. Most active surgical maneuvers in cataract surgery take place within the boundaries of the pupil since the cataract is anatomically located behind the iris and access to the cataract requires passage of instruments through the pupil. Medications, operating microscope illumination, and surgical maneuvers can alter the shape, size, and appearance of the pupil during different surgical phases.

A typical cataract surgery can be broken into 11 active surgical phases: Paracentesis, Medication and Viscoelastic Insertion, Main Wound, Capsulorrhexis Initiation, Capsulorrhexis Formation, Hydrodissection, Phacoemulsification, Cortical Removal, Lens Insertion, Viscoelastic Removal, and Wound Closure.^2^ These surgical steps involve different instrumentation and result in varying appearances of the pupil intraoperatively.

In the past decades, many studies have been proposed to investigate changes in the pupil related to cataract surgery.^3-9^ Ordiñaga-Monreal *et al.* recently investigated pupil diameters of 109 randomized eyes preoperatively and 3-months postoperatively using pupillometer software of the Topolyzer Vario.^5^ This group found that pupil size was reduced after cataract surgery, but also saw that the reduction was larger in men than in women. Ba-Ali *et al*. sought to evaluate the postoperative trajectory of pupil changes for patients undergoing cataract surgery.^7^ Maximal pupil diameter reduction was observed 3 weeks postoperatively, but subsequently recovered 3 months after surgery. The relation of cataract surgery to pupil size was also investigated by Rickmann *et al*.^8^ In that study, the pupil size of healthy participants was measured with the infrared-video PupilX pupillometer at different illumination levels before and after cataract surgery. This group also saw that pupil diameter decreased after cataract surgery but increased back to preoperative levels four weeks after cataract surgery.

Prior work examining pupillary changes related to cataract surgery has primarily focused on changes between *preoperative* and *postoperative* measurements of the pupil, ignoring *intraoperative* changes. Furthermore, most studies of surgery-related pupillary changes have relied on specialized pupillometry hardware not suitable for intraoperative use. Even when intraoperative pupil measurements have been obtained, they have been obtained manually and for very few time points.^9^

Previous studies demonstrated that an adequately dilated pupil is a prerequisite for safe cataract extraction.^10-13^ Vision-threatening complications of cataract surgery have been shown to be associated with pupillary instability.^14^ The automated analysis of intraoperative pupil changes could help surgeons perform large-scale studies to better understand risk factors for pupillary instability without the need for manual measurements that disrupt surgical flow and increase surgical time.

Pupillary dynamics are important beyond cataract surgery, and they affect the execution of vitreoretinal and corneal surgeries as well. In Descemet membrane endothelial keratoplasty (DMEK), for example, the timing of pupillary dilation and miosis plays an important role. Early dilation assists in retroillumination of the endothelium (facilitating descemetorrhexis), while later miosis is vital for protecting graft endothelial cells. Thus, while the context of the present study is cataract surgery, it is likely that applications for intraoperative pupil analysis exist for other domains of ophthalmic surgery as well.

In this study, we propose and validate a novel computational framework to track and analyze changes in pupil morphology *during* cataract surgery. The proposed framework consists of three primary parts: feature extraction, anatomy segmentation, and obstruction detection/compensation. This approach is designed to address the primary difficulties encountered in a standard system for recognizing pupils. These challenges include: 1) the potential for interference or obstruction caused by surgical instruments, eyelids, drapes, and similar structures. 2) the cropping of the pupil due to decentration in the camera sensor’s field of view, and 3) variations in magnification. It is hoped that this framework will enable the large-scale study of pupillary changes in not only cataract surgery, but ophthalmic surgery in general. Such studies will be necessary to identify clinical and intraoperative risk factors for pupillary instability as well as to develop intraoperative early warning systems for surgeons.

## METHODS

### BigCat Dataset Collection

One hundred ninety high-definition video recordings of cataract surgeries performed by surgeons at University of Michigan’s Kellogg Eye Center were collected in the period from 2020 to 2023. The study was approved (HUM00160950) by the Michigan Medicine IRB (IRBMED) in May of 2019. To obtain the high-quality videos in the dataset, Zeiss high-definition one-chip imaging sensors which were integrated into Zeiss Lumera 700 microscopes were used for recording the cataract surgeries at 1,920×1,080 resolution and a frame rate of 30 frames per second.

Periods of inactivity prior to surgery and after completion of surgery were trimmed. Frames were extracted from the video stream at a frequency of 15 frames per second (FPS). This strategic down-sampling was implemented to reduce the overall time required for subsequent analyses while retaining essential temporal information. The extracted frames were resized to dimensions of 480×270 pixels using bilinear interpolation to maintain sufficient video frame quality while minimizing time consumption.

To generate a dataset for the development and testing of models for anatomy recognition and analysis, the surgical videos were then processed to extract two random frames from each of 11 surgical phases. Ground truth surgical phase annotations were performed by trained human annotators for all frames within all 190 videos. Randomized selection of frames by phase was performed to ensure that pupil obscuration by a variety of surgical instruments associated with different phases of surgery was adequately represented in the dataset. In addition, since the Medication and Viscoelastic Insertion, Phacoemulsification, and Viscoelastic Removal phases have highly varied appearances (due to transient pupil distortion from viscoelastic instillation, nucleus disassembly, and eye rotation, respectively), an additional 2 frames per phase were randomly selected for these three phases. We further expanded the dataset by incorporating two additional frames from each video during which no active surgical maneuvers were being performed, termed a “No Activity” phase. The resulting dataset consisted of 5,700 frames that were resized to 480×270 pixels and stored in 24-bit Portable Network Graphics (PNG) format without editing. The number of images corresponding to each surgical phase in the collected dataset is shown in Table I.

**TABLE I.** Number of images included from bigcat by surgical phase.

| Surgical Phases | Number of Images |
| --- | --- |
| No Activity | 380 |
| Paracentesis | 380 |
| Medication and Viscoelastic Insertion | 760 |
| Main Wound | 380 |
| Capsulorrhexis Initiation | 380 |
| Capsulorrhexis Formation | 380 |
| Hydrodissection | 380 |
| Phacoemulsification | 760 |
| Cortical Removal | 380 |
| Lens Insertion | 380 |
| Viscoelastic Removal | 760 |
| Wound Closure | 380 |

Ground truth manual segmentations of anatomical components of the eye – the palpebral fissure, limbus, and pupil – were performed manually on all images in the dataset by trained human annotators. This annotation process was conducted using MATLAB version R2022a (The MathWorks; Natick, MA, USA) in conjunction with a Wacom One drawing tablet (Wacom Co., Ltd.; Kazo, Saitama, Japan) to ensure the accuracy of the annotations. The videos within the dataset were randomly divided into training, validation, and testing subsets consisting of 60%, 20%, and 20% of the dataset, respectively. As a result, the training set consisted of 3,420 images from 114 surgical videos, the validation set comprised 1,140 images from 38 videos, and the remaining 1,140 images from 38 videos were allocated for the testing set.

### Data Preprocessing and Feature Extraction

Significant variations in pupil appearance can occur during cataract surgery due to the presence of surgical instruments, hydration of the crystalline lens material, nuclear disassembly, and eventual replacement of the crystalline lens with the intraocular lens implant. Accordingly, even deep learning models can benefit from the utilization of tailored feature extraction methods for the task of semantic segmentation.^15^ In order to overcome these challenges, we propose the incorporation of an image preprocessing method in this phase to extract meaningful image features and attempt to improve downstream segmentation performance. A schematic of the overall framework is depicted in Fig. 1.

**Fig. 1.**
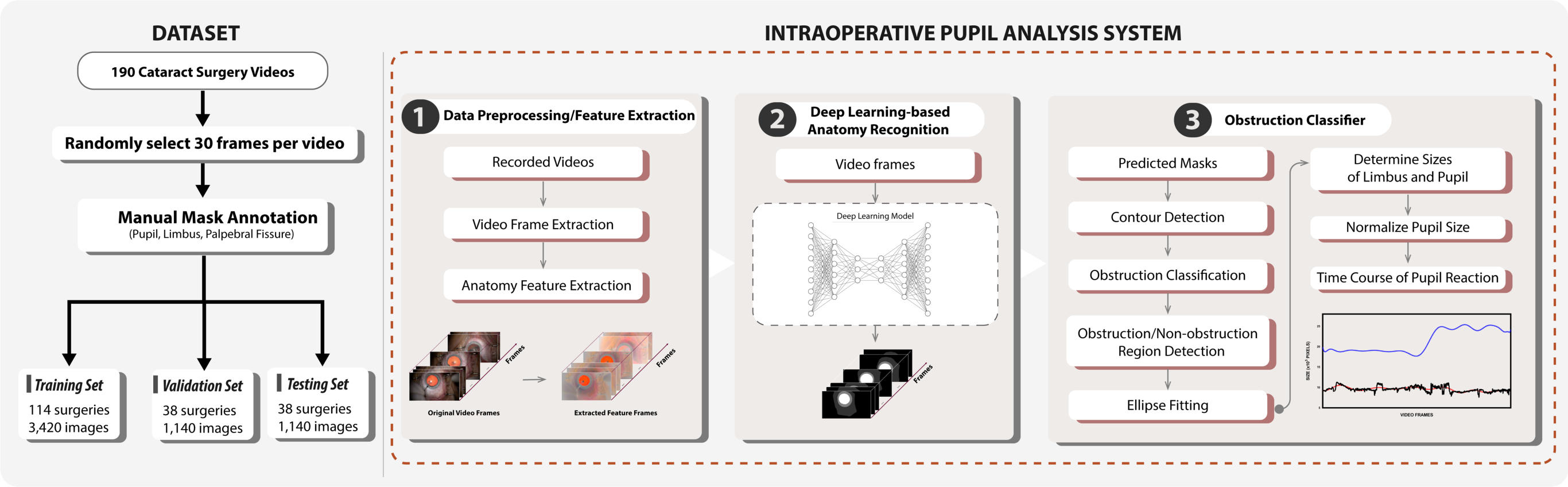
The proposed intraoperative pupil analysis framework for cataract surgery.

In this study, we employ the adaptive wavelet tensor feature extraction (AWTFE) method,^16^ described previously by our research group, for feature extraction. The primary objectives in employing the AWTFE method at this stage are: 1) to eliminate irrelevant information in the context of segmentation tasks, and 2) to extract and enhance the distinctive features of the pupil, limbus, and palpebral fissure within the image. The AWTFE method emphasizes object boundaries, textures, shapes, and other distinctive attributes relevant to the segmentation of the anatomical landmarks of interest. Sample output of the AWTFE method is depicted in Fig. 2. Evaluation of the impact of the AWTFE method on anatomy segmentation performance is described in the following section.

**Fig. 2.**
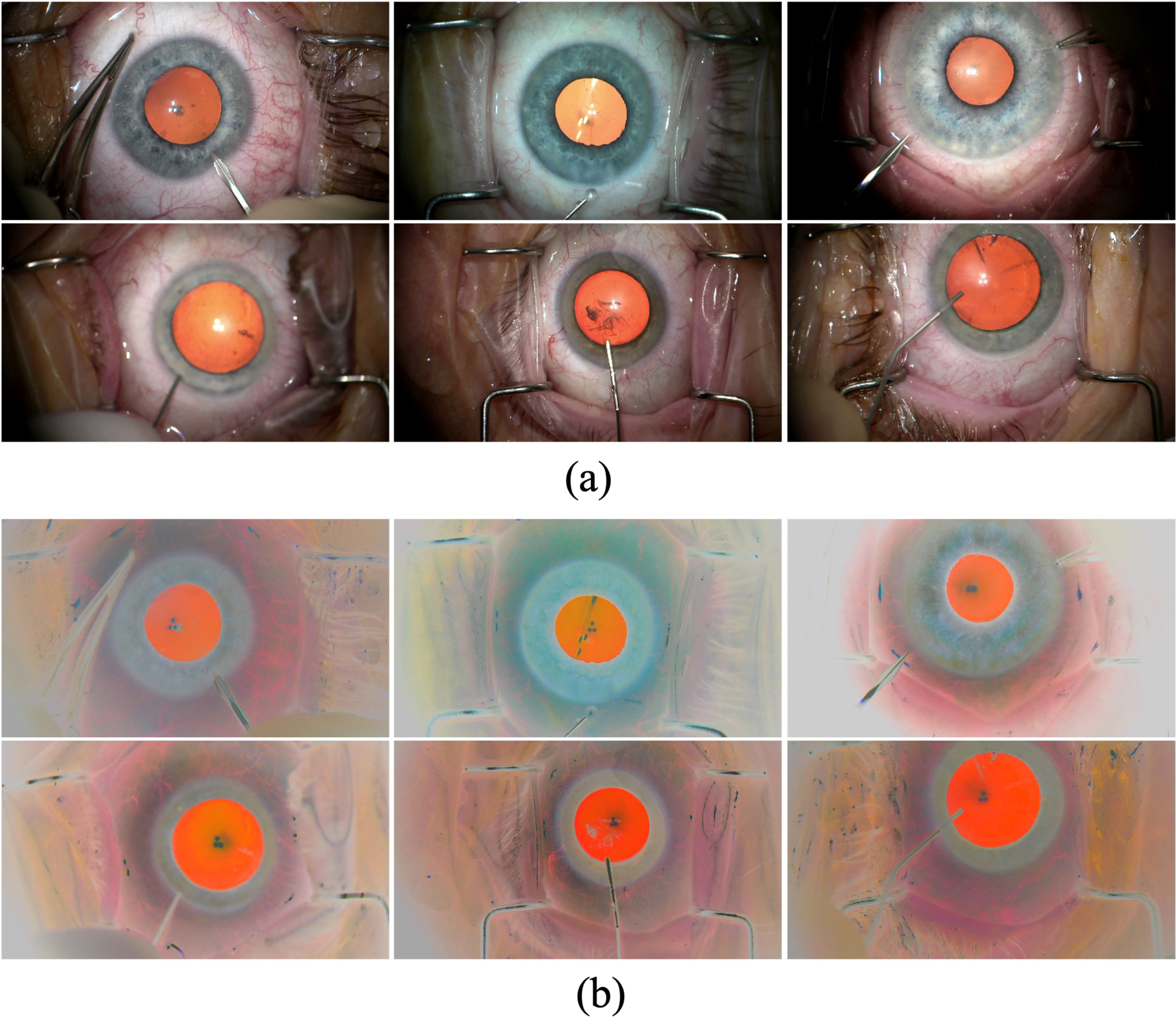
Anatomic feature extraction using the AWTFE method. (a) Original cataract surgery images in the dataset; and (b) Corresponding feature-extracted images generated by the AWTFE method.

### Deep Learning-based Anatomy Recognition

In the proposed framework, the accuracy of the analysis results heavily relies on anatomy segmentation. In this stage of the framework, the pupil, limbus, and palpebral fissure, are segmented within the video frames. Segmentation of the limbus and palpebral fissure was necessary for obstruction detection and compensation, as described in the next section. Three state-of-the-art deep learning-based segmentation models (UNet,^17^ LinkNet,^18^ and FPN^19^) and four distinct convolutional backbone networks (VGG16,^20^ ResNet50,^21^ DenseNet169,^22^ and MobileNet^23^) were considered to select an optimal combination for the anatomy segmentation task. Accordingly, 12 model-backbone combinations were studied in total. Backbone networks were each pre-trained on the ImageNet dataset.^24^ Two instances of each model-backbone combination were considered, one utilizing the AWTFE method for preprocessing of images, and one using raw input images. Accordingly, 24 models were studied in total.

The training set sizes were equivalent for all 24 models studied, with or without utilizing the AWTFE method. Training was performed with batch size of 32. All images were downsampled to 224×224 pixels for the input to the segmentation models. The Adam algorithm with an initial learning rate of 0.0001 was used as the optimizer for the segmentation models. The learning rate was then decreased by 0.1 when the validation loss stopped improving. Each model was trained for a maximum of 200 epochs, corresponding to 21,400 training iterations. Early stopping was implemented to avoid overfitting. The trained weights achieving the lowest validation loss during the training process were saved and utilized for validation.

In this study, the performance of the segmentation models was evaluated with precision and recall as the primary metrics, which are determined as follows:

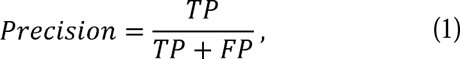

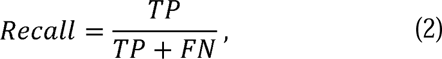

where *TP, FP, TN*, and *FN* denote the true positive, false positive, true negative, and false negative rates, respectively. In addition, the performance of the models was also evaluated by using the Intersection Over Union (IoU) and Dice Coefficient, which are determined as follows:

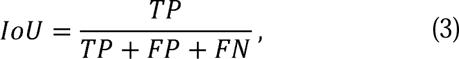

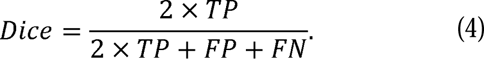

### Obstruction Detection and Compensation

The quantitative assessment of intraoperative pupil morphologic changes in cataract surgery presents challenges despite the ability to utilize deep learning models for semantic segmentation of the pupil region. In particular, three primary challenges are: 1) the potential for interference or obstruction caused by surgical instruments, eyelids, drapes, and similar structures. 2) the cropping of the pupil due to decentration in the camera sensor’s field of view, and 3) variations in magnification, as shown in Figs. 3(a) and (b). To address those challenges, our proposed framework introduces an Obstruction Detection and Compensation algorithm. This algorithm relies on automated segmentations of the pupil, palpebral fissure, and limbus generated during the segmentation phase to identify and compensate for obstructions. Furthermore, to ensure accuracy across diverse magnification settings, the computed pupil size is ultimately normalized by utilizing the predicted limbus size.

**Fig. 3.**
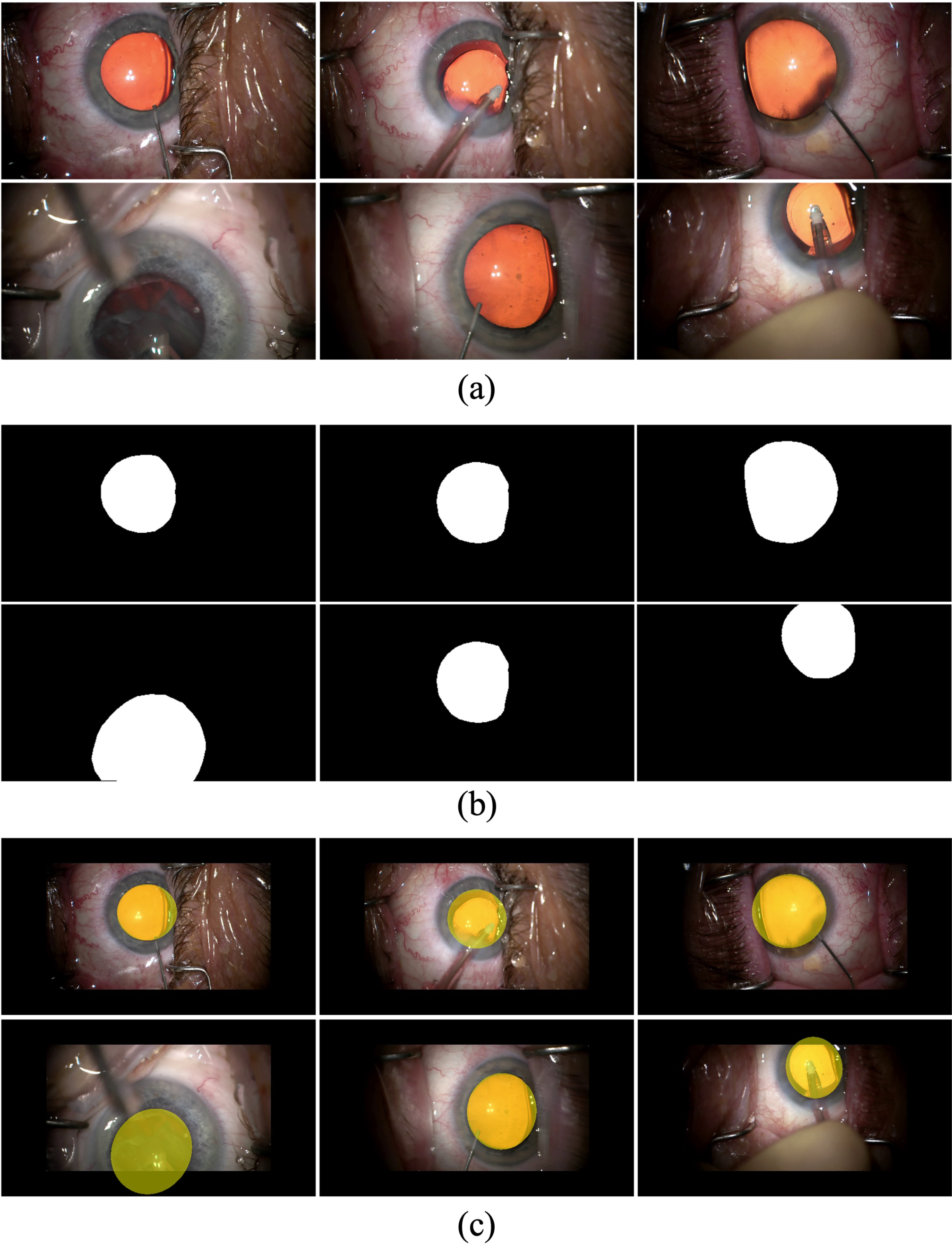
Sample obstruction compensation results. (a) Obstruction and decentration video frames; (b) Pupil segmentation masks generated by the deep learning model; and (c) Overlay images of pupil masks generated by the obstruction compensation algorithm.

The obstruction classifier utilizes a pair of masks generated by the deep learning models for the palpebral fissure and pupil, denoted as *M*_*q*_ and *M*_*p*_, respectively, to ascertain the presence of obstruction within a given frame. Initially, contours of both the palpebral fissure and the pupil are determined by binarizing the respective masks and applying edge computation through a Canny filter,^25^ as illustrated in Fig. 4. Subsequently, points *q*_*i*_ (*x*_*q*_, *y*_*q*_J and *p*_*j*_ (*x*_*p*_,*y*_*p*_J along the contours of the palpebral fissure and pupil are vectorized to enable the calculation of the Euclidean distance, *D*_*i,j*_, between these two vectors as follows:

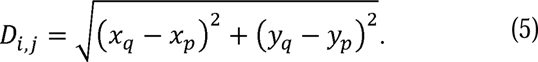

**Fig. 4.**
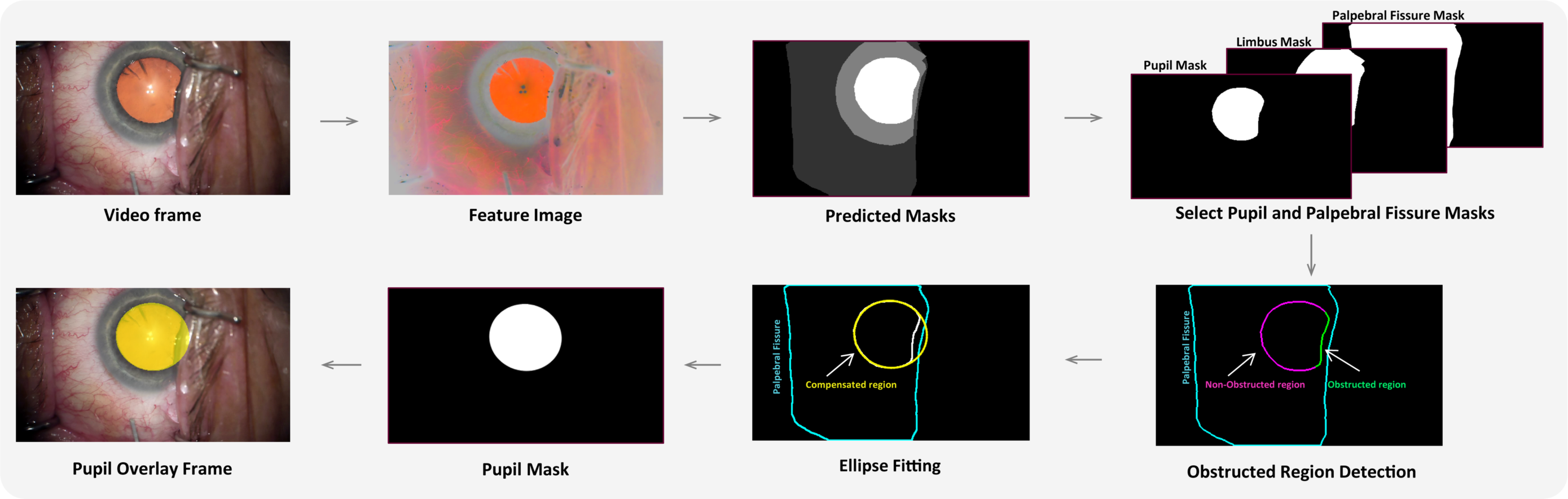
Schematic depicting the obstruction classifier embedded in the proposed pupil analysis framework.

The smallest computed Euclidean distance value is then compared against an obstruction threshold, *τ*. If the distance is larger than *τ*, the pupil region is devoid of obstruction, and the pupil size can be simply determined via the original pupil mask *M*_*p*_ generated by the deep learning model. In contrast, if the pupil is identified as an obstructed region based on the obstruction threshold *τ*, the subsequent steps involve determining obstructed and unobstructed segments. This is accomplished by calculating the minimum distance between each pupil point *p*_*j*_ (*x*_*p*_,*y*_*p*_J and all palpebral fissure points *q*_*i*_ (*x*_*q*_, *y*_*q*_J. If the minimum value is larger than *τ*, *p*_*j*_(*x*_*p*_,*y*_*p*_J is treated as an unobstructed point, and vice versa. Subsequently, an ellipse, which is considered an estimate of the pupil region, is fitted using the unobstructed points.^26^ By considering solely the unobstructed pupil points, the estimated ellipse eliminates noise caused by obstructed points on the pupillary boundary. To this end, the estimated pupil size P can be established by computing the area of the fitted ellipse. The results of this process are shown in Fig. 3(c).

The magnification of images within a recorded video may vary due to operating microscope adjustments made by surgeons during surgery. To address this challenge, the pupil size, which was previously estimated as above, was normalized using the corresponding size of the (obstruction-compensated) limbus region. In contrast to the pupil, the size of the limbus region is physically fixed and cannot be significantly altered under normal circumstances during phacoemulsification. However, the size of the segmented limbus can also be affected by inaccuracies resulting from surgical obstructions. Therefore, to ensure the accuracy of the system, the actual size of the limbus, denoted as *L*, is first estimated by the obstruction compensation algorithm described above using the predicted masks of the palpebral fissure and limbus. The size of the pupil region at *k*_*th*_ frame *P*_*k*_ is then normalized with respect to the size of the corresponding limbus region *L*_*k*_ as follows:

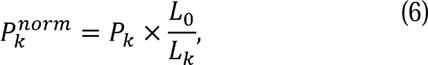

where *L*_0_ is determined as the limbus region computed directly from the predicted mask of the first frame by the deep learning model. This ensures that the computed size of the pupil region at *k*_*th*_ frame *P*_*k*_ is adjusted for any magnification changes made intraoperatively.

In order to assess the effectiveness of this approach, we evaluated the performance of the proposed framework in its capability to classify obstructed frames and estimate the actual size of the pupil using 1,140 images selected randomly from 38 videos in the test set. The images were manually classified into pupil obstruction (*OB*) and non-obstruction (*NOB*) classes by two trained annotators. Accordingly, 22.45% of the testing set (256 images) belonged to the *OB* class and 77.55% of the testing set (884 images) were in *NOB* class. Furthermore, the actual pupil region of the images in the *OB* class was manually estimated and annotated to be used as the ground truth for the experiment.

### Downstream Analysis Example: Prediction of Pupil Expansion Device Use

In order to examine the utility of the output of the pupil analysis framework described above, we investigated whether the timecourse of pupillary area was predictive of surgeon usage of a pupil expansion device (PED) in a separate dataset.

The use of a PED by an experienced surgeon is an indication that the surgeon detects pupillary characteristics that may impair successful completion of the surgery. A PED is preferably placed prior to the creation of the anterior capsulotomy, so as to avoid inadvertently capturing the capsulotomy edge with the PED. Accordingly, we investigated whether the pupil area time series generated by our framework for the surgical phases prior to the initiation of the capsulorrhexis (excluding PED placement in those cases that involved PED placement) could be used to predict whether a PED would later be placed.

We first constructed a dataset that included the mean pupil sizes during three early surgical phases of cataract surgery: Paracentesis, Medication and Viscoelastic Insertion, and Main Wound, across 50 cases with PED placement and 50 cases without PED placement. In order to identify the surgical phase within PED videos, we employed the CatStep surgical phase classification model.^25^ It is noted that the CatStep model utilized was not previously trained on PED videos. Hence, we conducted manual validation of its phase classification results to ensure the accuracy of phase boundaries in the PED videos. The videos in the dataset were randomly divided into training (70%) and testing (30%) sets, maintaining the balance of classes in the training and testing sets. We investigated the capacity of classification models, including Support Vector Machine (SVM),^27^ K-Nearest Neighbors (KNN),^28^ Random Forest,^29^ Decision Tree,^30^ Naïve Bayes,^31^ and Logistic Regression,^32^ to predict PED usage based on the averaged pupil size time series. All experiments in this study were implemented on a workstation with a 24-Core Xeon Intel CPU, with 128 GB RAM and four NVIVIA RTX 2080 Ti GPUs running Ubuntu.

## RESULTS

In this section, the performance of the proposed framework is presented in detail for each of its components. Additionally, we present an analysis of pupillary changes by phase of surgery. To demonstrate the potential for downstream analyses utilizing the pupil analysis framework, we also evaluate the performance of algorithms for predicting PED use based on the pupillary size timecourse derived from the framework.

### Feature Extraction and Anatomy Segmentation

The anatomy segmentation performance of the deep learning models considered is detailed in Table II. The FPN architecture combined with the VGG16 backbone demonstrated the highest mean Dice coefficient (95.03%) across the three anatomic segmentation classes (pupil, limbus, and palpebral fissure) compared to all other model-backbone combinations.

**TABLE II.** Segmentation performance of deep learning models with and without the awtfe method on the validation set.

| NETWORK ARCHITECTURE | Backbone Network | Precision (%) | Recall (%) | IoU (%) | Dice (%) |
| --- | --- | --- | --- | --- | --- |
| UNet | MobileNet | 94.09 | 94.69 | 89.52 | 94.15 |
| UNet + AWTFE |  | **95.86** | **95.89** | **92.28** | **95.66** |
| LinkNet |  | 93.47 | 93.90 | 88.27 | 93.14 |
| LinkNet + AWTFE |  | **95.52** | **95.49** | **91.63** | **95.27** |
| FPN |  | 93.53 | 93.97 | 88.37 | 93.46 |
| FPN + AWTFE |  | **95.67** | **96.33** | **92.45** | **95.79** |
| UNet | DenseNet169 | 94.72 | 94.92 | 90.28 | 94.63 |
| UNet + AWTFE |  | **96.47** | **96.57** | **93.45** | **96.39** |
| LinkNet |  | 94.43 | 94.60 | 89.74 | 94.31 |
| LinkNet + AWTFE |  | **96.08** | **96.44** | **92.99** | **96.13** |
| FPN |  | 94.20 | 94.78 | 89.68 | 94.28 |
| FPN + AWTFE |  | **96.39** | **96.49** | **93.26** | **96.31** |
| UNet | ResNet50 | 94.86 | 95.12 | 90.57 | 94.77 |
| UNet + AWTFE |  | **96.64** | **96.49** | **93.49** | **96.41** |
| LinkNet |  | 94.35 | 94.68 | 89.71 | 94.28 |
| LinkNet + AWTFE |  | **95.99** | **96.63** | **93.04** | **96.14** |
| FPN |  | 94.61 | 94.79 | 90.06 | 94.47 |
| FPN + AWTFE |  | **96.36** | **95.99** | **92.79** | **95.98** |
| UNet | VGG16 | 94.96 | 95.04 | 90.58 | 94.79 |
| UNet + AWTFE |  | **96.63** | **96.03** | **93.11** | **96.18** |
| LinkNet |  | 94.25 | 94.65 | 89.63 | 94.19 |
| LinkNet + AWTFE |  | **96.33** | **96.07** | **92.85** | **95.99** |
| FPN |  | 94.99 | 95.45 | 91.00 | 95.03 |
| FPN + AWTFE |  | **96.41** | **96.87** | **93.66** | **96.52** |

Bold fonts indicate the better performance across models

The performance of every model-backbone combination was improved through the addition of feature extraction using the AWTFE method (p < 0.0001). Mean Dice coefficients across the three segmentation classes of all investigated models were improved by up to 2.23%. The FPN-VGG16 architecture with AWTFE feature extraction outperformed all other models considered, with a Dice coefficient of 96.52%. The FPN-VGG16+AWTFE model outperformed the original FPN-VGG16 network by 1.49%. Accordingly, the FPN-VGG16+AWTFE model was selected for incorporation into the pupil analysis framework.

### Obstruction Detection and Compensation Performance

In order to optimize the performance of the obstruction detection component of the proposed framework, we first examined the impact of threshold values, *τ*_*P*_, on the accuracy of the obstruction classifier for pupil on the 1,140 images in the testing set. As shown in Fig. 5, the highest classification performance (*Dice*) was obtained with *τ* = 8. With this value of *τ*, the edge detection-based classifier achieved a Dice coefficient of 75.74%. The classification performance gradually decreased with increasing values of *τ*. By using the proposed framework with *τ* = 8, with obstruction compensation applied as described in the Methods section, a Dice coefficient of 96.82% for pupil segmentation was achieved across the 1,140 test set images.

**Fig. 5.**
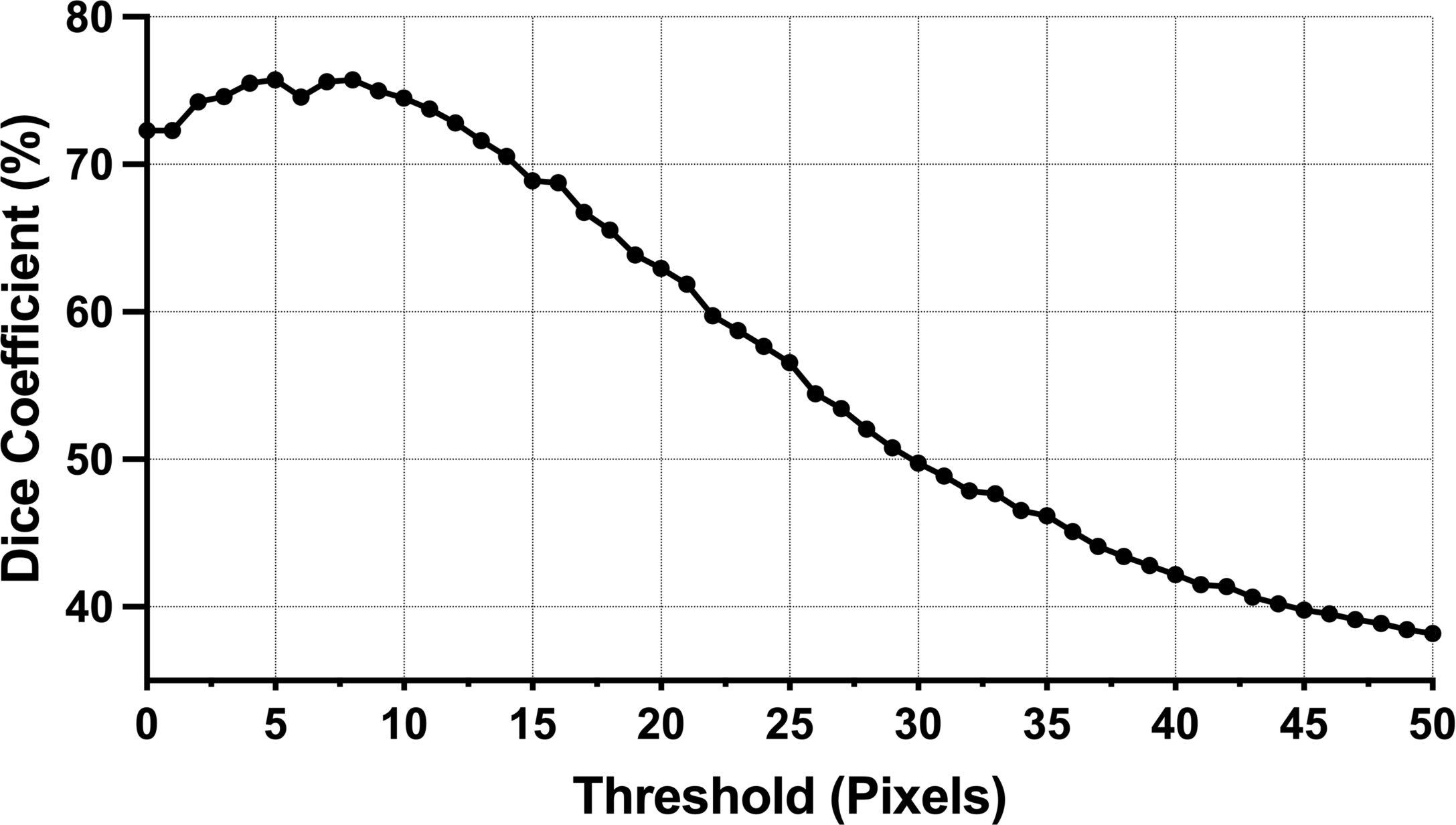
Classification performance of the obstruction classifier on 1,140 images of 38 surgical videos.

Utilization of obstruction detection led to an increase in pupil segmentation performance by 0.51%, as shown in Fig. 6(a). The Dice coefficients for all 11 active phases of surgery were significantly higher with obstruction detection and compensation than without obstruction detection (p = 0.0002).

**Fig. 6.**
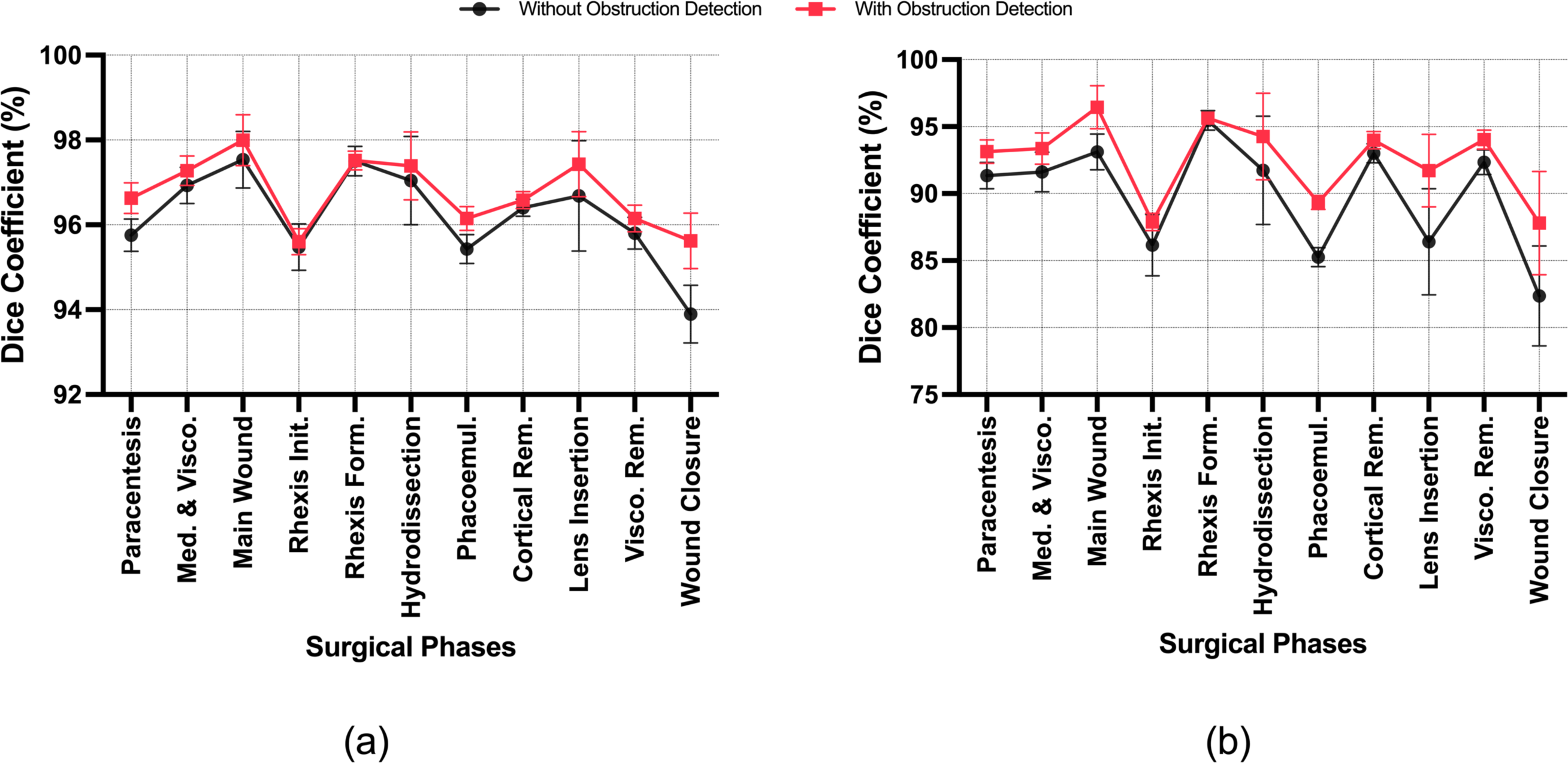
Dice coefficients by surgical phase for the proposed framework with and without obstruction detection. (a) The results on all 1,140 images in the testing set and (b) The results on all 256 obstruction images in the testing set. (Med. & Visco. = Medication and Viscoelastic Insertion; Rhexis Init. = Capsulorrhexis Initiation; Rhexis Form. = Capsulorrhexis Formation; Phacoemul. = Phacoemulsification; Cortical Rem. = Cortical Removal; Visco. Rem. = Viscoelastic Removal).

Of the 1,140 images in the test set, 256 (22.45%) had obstructions (*OB* class), while the remaining 884 did not (*NOB* class). To evaluate more directly the impact of the obstruction detection and compensation system when obstructions were present, performance was assessed further on the *OB* subset. The framework with obstruction detection and compensation achieved a Dice coefficient of 93.12%, whereas the framework without obstruction detection yielded only 90.88% (p = 0.0014J. Phase-wise performance differences in the *OB* subset are depicted in Fig. 6(b).

The processing time required by the proposed analysis system for an obstructed frame was approximately 19.10 milliseconds (ms) from start to finish. Of the 19.10 ms, the obstruction detection and compensation component required only 4.30 ms. These findings reveal that the proposed system is well-suited for real-time intraoperative applications, with a throughput of over 52 FPS while maintaining the accuracy described above.

### Phase-Based Pupil Reaction Analysis

To analyze phase-based changes in pupil size, we randomly selected 15 videos from the testing set (none determined to have intraoperative floppy iris syndrome and none requiring a pupil expansion device) and executed the framework on the entire videos. The pupil size timecourses for four surgical cases analyzed using the proposed framework are depicted in Fig. 7. The mean pupil sizes within the 11 active surgical phases relative to the initial pupil size determined at the beginning of each surgery are shown in Fig. 8. An increase in pupil size is seen following the Medication Injection (buffered lidocaine and epinephrine) and Viscoelastic Insertion. During this phase, the pupil size increased by an average of 9.64% compared to the initial pupil size.

**Fig. 7.**
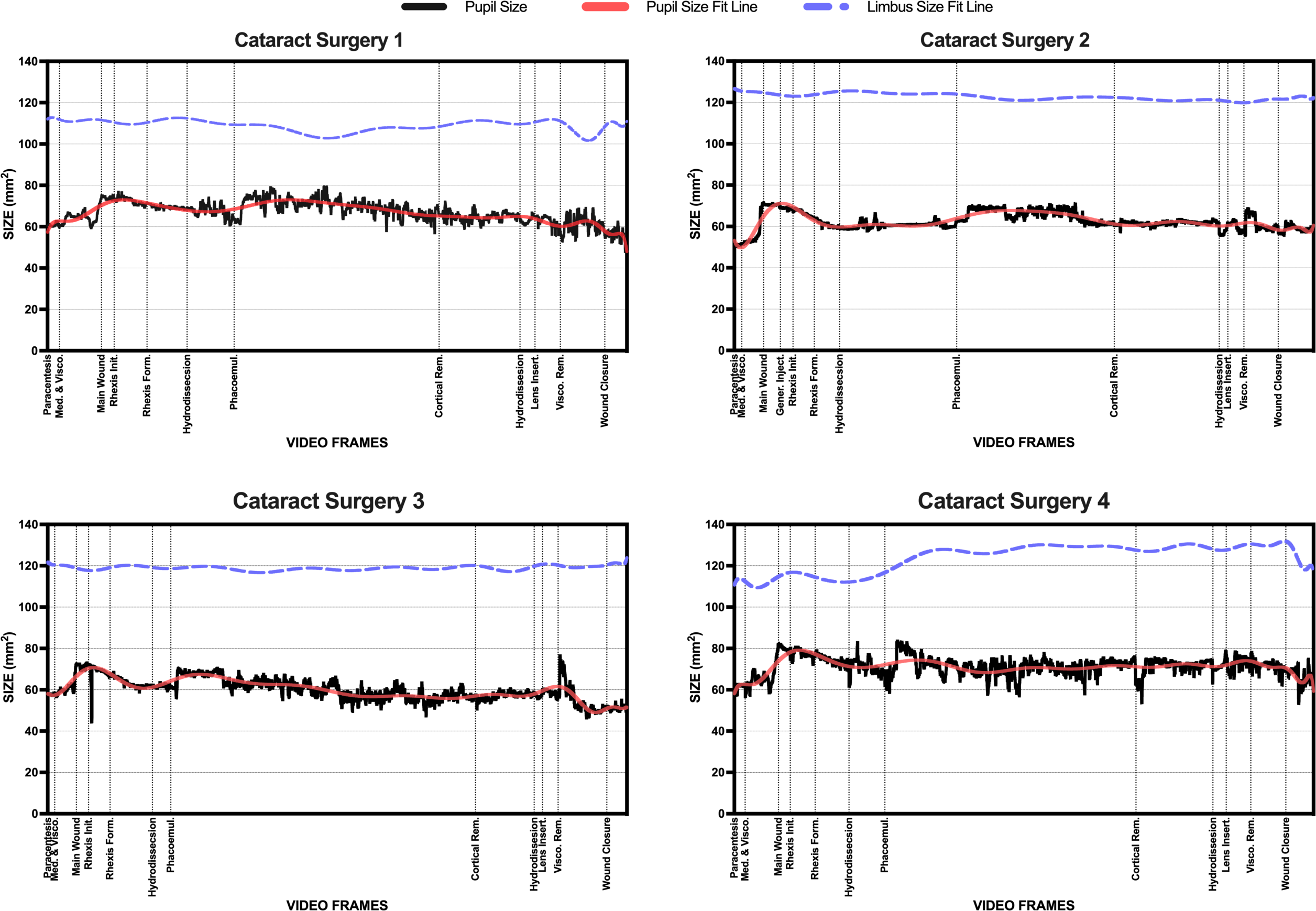
Pupil and limbus area output from the proposed framework for four cataract surgery cases with phases of surgery indicated. (Med. & Visco. = Medication and Viscoelastic Insertion; Rhexis Init. = Capsulorrhexis Initiation; Rhexis Form. = Capsulorrhexis Formation; Phacoemul. = Phacoemulsification; Cortical Rem. = Cortical Removal; Visco. Rem. = Viscoelastic Removal).

**Fig. 8.**
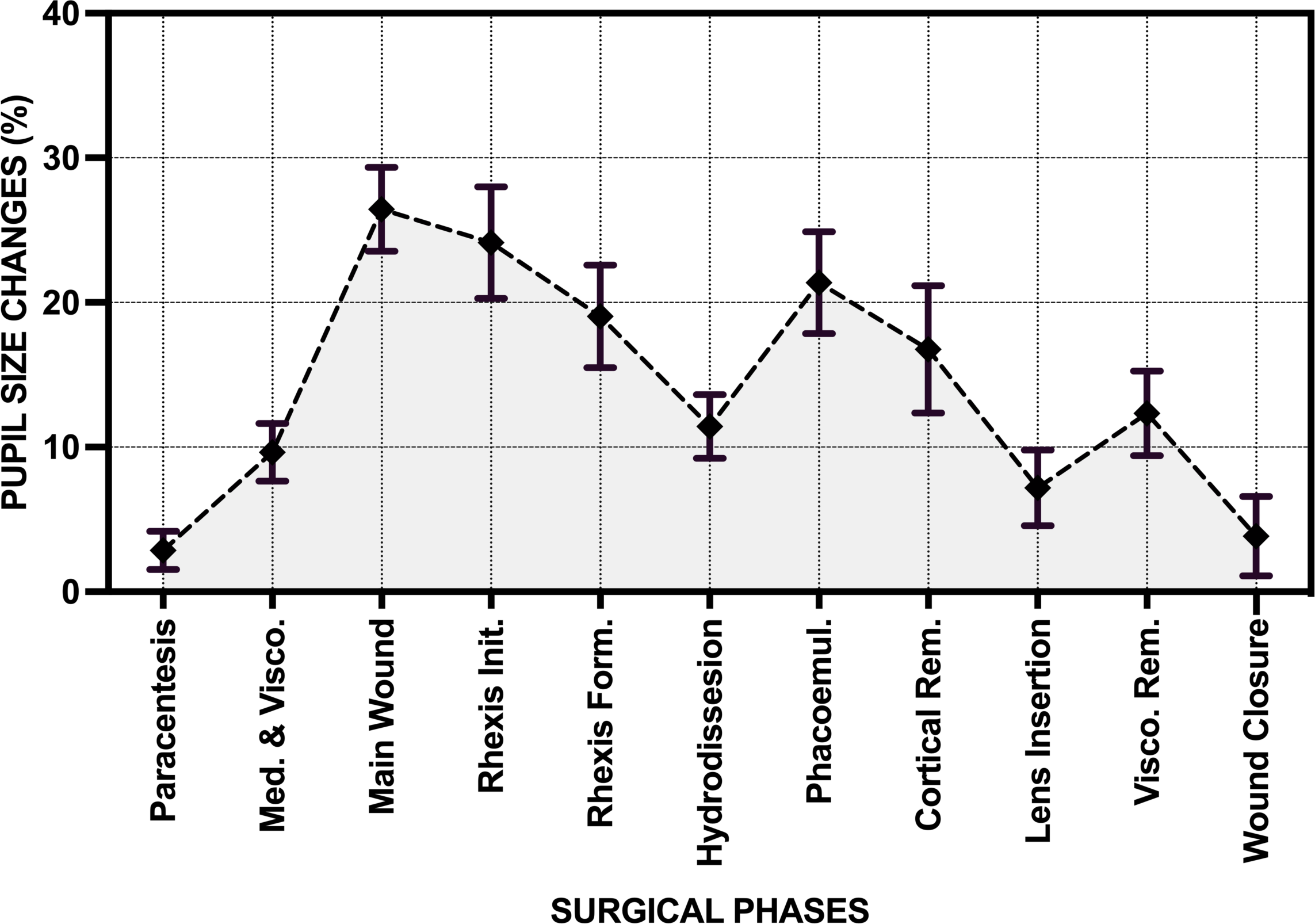
Mean pupil size by surgical phase relative to preoperative pupil size across 15 surgical videos. (Med. & Visco. = Medication and Viscoelastic Insertion; Rhexis Init. = Capsulorrhexis Initiation; Rhexis Form. = Capsulorrhexis Formation; Phacoemul. = Phacoemulsification; Cortical Rem. = Cortical Removal; Visco. Rem. = Viscoelastic Removal).

The Main Wound phase was consistently followed by a reduction in pupil size, a pattern evident in all trajectories plotted in Fig. 7. This phase shows the highest mean pupil size, which then gradually decreased in subsequent phases, including Capsulorrhexis Initiation, Capsulorrhexis Formation, and Hydrodissection, as detailed in Fig. 8. The reduction in pupil size may be related to loss of viscoelastic through the main wound during these phases. Ultimately, the pupil size post-Wound Closure closely approximated the preoperative pupil size (higher by just 0.99% relative to the initial size).

### Pupil Expansion Device Use Prediction Performance

In order to evaluate the utility of the pupil size timecourse output by the proposed framework for downstream analyses, attention was then turned to a dataset of 50 videos with PED placement and 50 videos without PED placement (as described in the Methods section). In comparison to standard surgeries performed without PED, pupil size during the initial phases (Paracentesis, Medication and Viscoelastic Insertion, and Main Wound) of PED surgeries were not significantly different (p = 0.375).

Six classification models were trained using pupil size timecourses from the dataset of 50 PED and 50 non-PED videos. Only the timecourses from the Paracentesis, Medication and Viscoelastic Insertion, and Main Wound phases were included for this analysis in order to simulate surgeon decision-making. In our experiment, all classification algorithms were trained on the training set consisting of the pupil size timecourses of 70 surgeries and validated on the testing set, which included 30 surgeries. The performance of the classification models on the testing set is shown in Table III and Fig. 9. Among the six machine learning models, the Random Forest and Support Vector Machine (SVM) models achieved the highest classification accuracy at 96.67% and the highest AUC (Area Under the Curve) of 99.33 and 99.44%, respectively. These experimental results demonstrate that the intraoperative pupil size across early surgical phases can be effectively utilized to predict surgeon PED usage in cataract surgery.

**Fig. 9.**
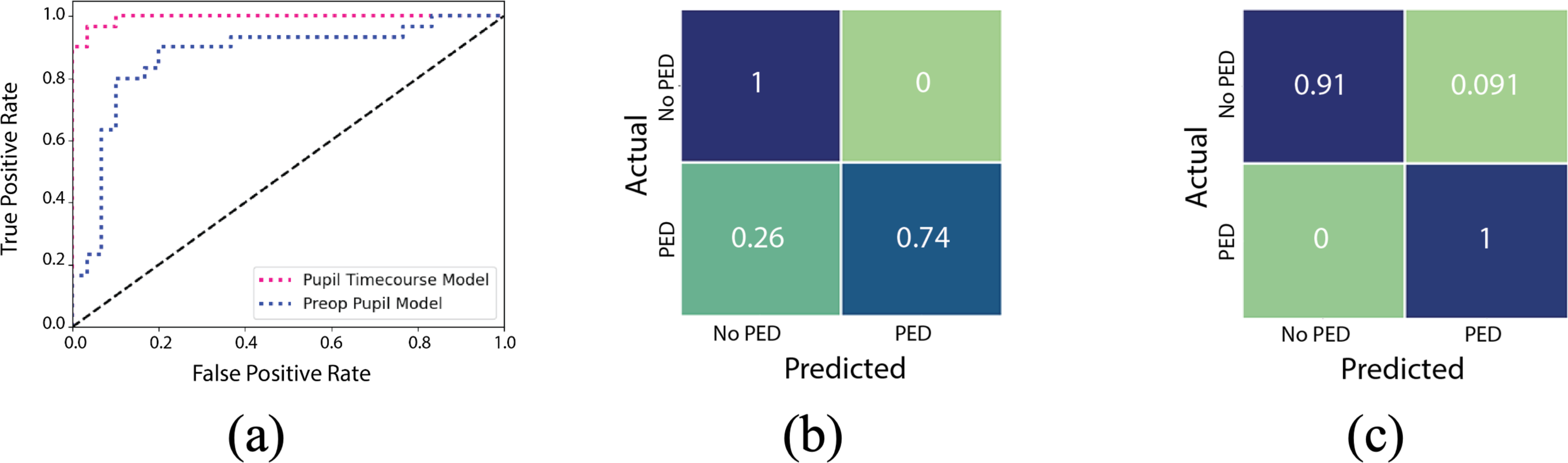
The performance comparison for prediction of pupil expansion device (PED) usage of the classification models using the pupil timecourses and the preoperative pupil size. (a) The Receiver Characteristic Curves (ROC) of the pupil timecourse model (SVM) and the preoperative pupil model (Logistic Regression); (b) The confusion matrix of the preoperative pupil model; and (c) The confusion matrix of the pupil timecourse model.

**TABLE III.** Pupil expansion device usage prediction performance of machine learning models.

| Models | Preoperative Pupil Size |  | Pupil Size Timecourse |  |
| --- | --- | --- | --- | --- |
|  | Accuracy (%) | AUC (%) | Accuracy (%) | AUC (%) |
| Naïve Bayes | 83.33 | 87.00 | **93.33** | **93.33** |
| KNN | 76.67 | 87.33 | **83.33** | **89.44** |
| Logistic Regression | 83.33 | 87.22 | **90.00** | **97.74** |
| Decision Tree | 73.33 | 73.33 | **93.33** | **93.33** |
| SVM | 83.33 | 87.00 | **96.67** | **99.44** |
| Random Forest | 73.33 | 86.56 | **96.67** | **99.33** |

Bold fonts indicate the better performance

We then employed the mentioned machine learning models trained with preoperative pupil size data alone instead of using pupil size timecourses. Preoperative pupil size was determined at the initial frame of each surgical video using our proposed system. Our primary objective was to evaluate the effectiveness of using the timecourse generated by our proposed system compared to the traditional approach of relying on preoperative pupil size measurements performed by surgeons in clinic or at the beginning of surgery. The models trained on preoperative pupil size alone achieved lower performance compared to those trained on pupil size timecourses from the early surgical phases. Notably, Logistic Regression achieved the highest performance among the models trained on preoperative pupil sizes with an accuracy of 83.33% and an AUC of 87.22%, which are significantly lower than the highest performance of the machine learning model achieved on the pupil size timecourses by 16.34% and 12.22%, respectively. These results indicate that prediction of PED usage can be more performed more accurately using timecourses generated by proposed intraoperative pupil analysis framework than using preoperative pupil size alone.

## DISCUSSION

In the present study, we have proposed and validated a computational framework for intraoperative analysis of pupil morphology during cataract surgery. The system involves three primary stages: feature extraction, anatomy segmentation, and obstruction detection/compensation. In the feature extraction stage, we employed the AWTFE method, previously developed by our group, to generate feature-rich versions of video frames for the effective segmentation of relevant anatomical structures. When combined with state-of-the-art deep learning-based segmentation models, the AWTFE method significantly improved segmentation performance for all models considered. The FPN model with VGG16 backbone and AWTFE feature extraction (FPN-VGG16+AWTFE) was found to have the highest performance in validation and was ultimately chosen for incorporation into the framework. The final model achieved a Dice coefficient of 96.52% when evaluated on the held-out testing set.

In order to ensure the reliability of the system in delivering precise intraoperative analysis of pupillary changes, we introduced a novel obstruction detection/compensation component. This innovation addresses typical issues arising from obstructions and varying image scales in cataract surgery videos. Our findings demonstrate that when employing the proposed obstruction detection and compensation algorithm, the system can achieve an overall Dice coefficient of 96.82% for pupil segmentation and can improve segmentation performance by 2.24% for frames in which obstructions were present. Since pupil-obstructed frames comprised 22.45% of the randomly selected test set, obstruction detection and compensation are likely to be essential for ensuring the reliability of downstream time-series analyses of pupillary metrics.

The run-time assessment revealed that the proposed analysis system can fully process an obstructed frame in under 19.10 milliseconds, achieving throughput of over 52 frames per second (FPS). Since cataract surgeries involve a mix of obstructed and unobstructed frames, 52 FPS represents a lower bound on throughput for the proposed system running on the described hardware. The high throughput (greater than 30 FPS) and high accuracy of the proposed system indicate its potential for future real-time intraoperative use as part of a broader decision support or analysis system.

We utilized the proposed system to examine pupil trajectories through the various phases of cataract surgery. Findings such as 1) the increase in pupil size after medication and viscoelastic injection, 2) the reduction in pupil size during the course of phacoemulsification, and 3) the reduction in pupil size after viscoelastic removal align with typical surgeon experience.

To demonstrate the potential for downstream analysis of the pupil morphology timecourse generated by the proposed framework, we performed additional experiments to assess the ability to predict pupil expansion device (PED) usage using output from the framework. Using only pupil morphology data in the first three phases of surgery (Paracentesis, Medication and Viscoelastic Insertion, and Main Wound), it was possible to create an SVM classifier with 96.67% accuracy and 99.44% AUC in predicting the eventual use of a pupil expansion device during surgery. The performance of this approach was significantly higher than what could be achieved with preoperative pupil size data alone. The final pupil timecourse-based SVM could serve as part of a decision support system for trainees or early-stage attending surgeons and demonstrates the potential for building upon the output of our proposed framework.

Limitations of the study include the utilization of surgical videos from a single institution. However, the AWTFE method has previously been validated on the external CaDIS dataset, and the BigCat dataset utilized here is the largest database of deeply annotated surgical video in the world, comprising over 4 million frames in total. Furthermore, the proposed framework is seen as a starting point for intraoperative pupil analysis, since additional challenges remain. These challenges include compensation for ocular rotation, anterior chamber depth, and corneal power, which will be addressed in future studies.

Given the demonstrated performance contributions of each component of the computational framework proposed here, it appears this framework can serve as a reliable foundation for further analyses of intraoperative pupillary changes. We believe that the proposed framework can serve as a research tool for the large-scale examination of preoperative risk factors for pupillary instability, which is known to be associated with increased rates of complications of cataract surgery. While the current work has focused on cataract surgery, the framework is likely to be applicable to other pupil-sensitive forms of surgery, including vitreoretinal surgery and corneal surgery. Future work will attempt to validate this framework for other forms of ophthalmic surgery and explore additional downstream analyses and decision support scenarios.

## Data Availability

Data produced in the present study are not publicly available.

